# A data-driven system for insulin pump dosing in type 2 diabetes: development, prospective evaluation and clinical applicability

**DOI:** 10.64898/2026.10.13.26365400

**Authors:** Lewen Cai, Shuhong Huang, Yuping Cao, Zhimin Huang

## Abstract

Open-loop insulin pump therapy depends on clinical expertise for dose initiation and titration. We developed a long short-term memory-based system, using 1738 treatment courses from 1641 patients with type 2 diabetes for initial prescribing and daily basal and bolus titration. Patient-grouped five-fold validation showed closer agreement with recorded prescriptions for subsequent titration than for initiation. Prospective evaluation in 97 patients from the model-directed arm of a randomized trial yielded a median derived time in range (3.9–10.0mmol/L) of 84.5%, with low hypoglycemic exposure. Favorable control extended to previously treated patients. Higher mean glucose during the first 24 hours was associated with poorer glycemic control thereafter. The association between distance from development data and glycemic outcomes was stronger when early glucose responses were included alongside baseline characteristics. These findings support model-directed pump dosing and highlight early treatment response as a clinically relevant signal for assessing applicability and identifying patients who may require additional oversight. Trial registration: ClinicalTrials.gov, NCT07066891.

## 1 Introduction

Open-loop continuous subcutaneous insulin infusion (CSII) is widely used for hospital-based management of type 2 diabetes in China, supported in large part by basic medical insurance coverage for most inpatient pump-related costs. Its principal applications are short-term intensive insulin therapy (IIT) and inpatient glycemic management, including in non-endocrinology wards, as an alternative to multiple daily injections (MDI). Short-term IIT alleviates glucotoxicity and improves *β*-cell function and insulin sensitivity, facilitating long-term drug-free remission in a substantial proportion of patients with newly diagnosed diabetes, and potentially permitting deescalation to a simpler maintenance regimen in selected patients with longer-standing disease [1–5]. CSII allows flexible adjustment of basal rates and meal boluses without additional injections, facilitating more efficient dose titration than MDI. However, because open-loop pumps do not adjust insulin delivery autonomously, clinicians must integrate patient characteristics with evolving glucose profiles to determine initial and subsequent doses. This process depends heavily on clinical judgment and experience, contributing to variation in glycemic outcomes and hypoglycemia risk across clinicians and institutions. Suboptimal control during CSII may also limit subsequent drug-free remission or treatment simplification.

Data-driven artificial intelligence (AI) offers a means of learning the complex relationships among patient characteristics, longitudinal glucose trajectories, and insulin requirements. Recent clinical studies indicate that AI-assisted insulin titration can achieve glycemic efficacy and safety comparable to clinician-directed care, including care provided by experienced endocrinologists, while reducing the burden of repeated dose adjustments [6, 7]. These evaluations, however, have focused primarily on intermittent subcutaneous injection regimens administered one to four times daily. To our knowledge, no prospective study has evaluated a data-driven AI system integrating initial prescribing with subsequent daily basal–bolus titration during open-loop CSII in hospitalized patients with type 2 diabetes. The performance, robustness, and generalizability of data-driven models depend critically on the quality and representativeness of the data used for their development [8, 9]. Consequently, strong overall performance does not necessarily translate into equally reliable recommendations for patients who may be underrepresented in the development data. For a system intended to direct insulin dosing with minimal routine physician adjustment, it is therefore essential to assess its clinical applicability and identify circumstances that may warrant additional oversight [10, 11].

Building on more than two decades of clinical practice and research in CSII-based short-term IIT at the Department of Endocrinology, the First Affiliated Hospital of Sun Yat-sen University, the present study drew on a longitudinal dataset of patient characteristics, glucose measurements, and insulin pump prescriptions extracted from routine clinical records. This resource was used to develop a long short-term memory (LSTM)-based system that recommends insulin doses at CSII initiation and throughout subsequent daily titration. The system was intended to translate accumulated clinical expertise into a standardized and scalable dosing strategy with less reliance on highly experienced endocrinologists. This study reports the development and internal validation of the system, evaluates its prospective performance during model-directed treatment, and explores factors associated with suboptimal glycemic response (Fig. 1).

**Fig. 1.**
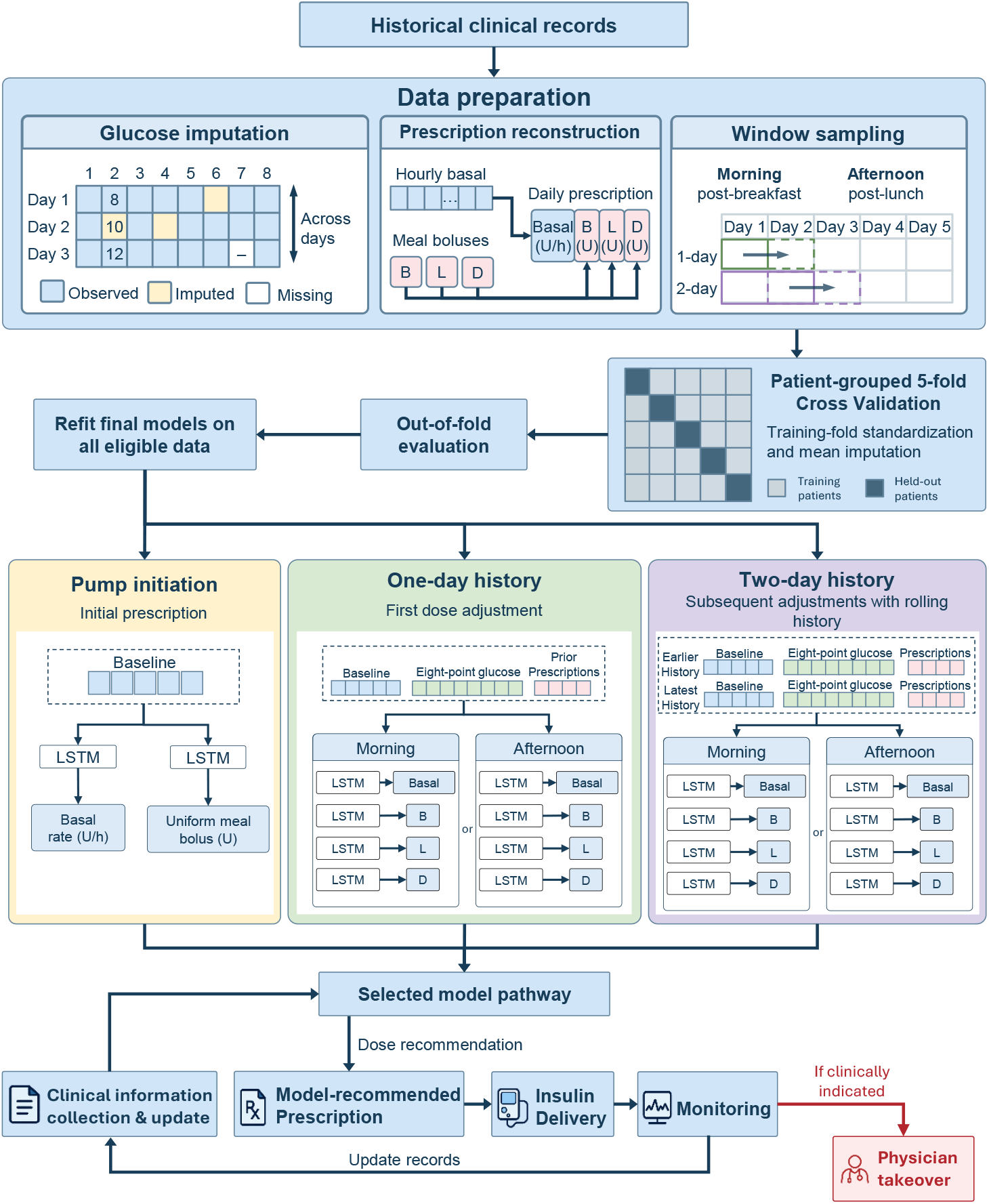
Overview of system development, internal validation and clinical implementation. Historical clinical records underwent glucose imputation, prescription reconstruction and sliding-window sampling. Patient-grouped five-fold cross validation generated out-of-fold (OOF) predictions for internal validation, followed by refitting on all eligible development data. Eighteen task-specific long short-term memory (LSTM) models supported continuous subcutaneous insulin infusion (CSII) dosing. Pump-initiation models used five baseline features, whereas subsequent models additionally incorporated glucose and prescription histories through morning or afternoon pathways. One-day history supported the first dose adjustment after pump initiation, and rolling two-day history supported subsequent adjustments. Model recommendations were issued as prescriptions by physicians, with updated clinical records informing subsequent recommendations and physician takeover when clinically indicated. B, L and D denote breakfast, lunch and dinner boluses, respectively.

## 2 Results

### Composition of the model-development dataset

A total of 1641 unique patients with newly diagnosed type 2 diabetes contributed 1738 eligible CSII treatment courses, comprising 25027 treatment days in this study. Most patients underwent one treatment course (n = 1558), whereas 72 and 11 patients underwent two and three or more courses, respectively. Among patients who underwent more than one course, 97 intervals between successive courses were identified, 92 of which were documented periods without glucose-lowering medication. These medication-free intervals ranged from 0.3 to 12.3 years, with a median duration of 3.6 years (IQR, 2.2-5.7).

The model-development dataset predominantly represented middle-aged patients, with male patients contributing 68% of treatment courses. Hyperglycemia before CSII initiation was marked, with a mean HbA1c of 10.9% and a mean fasting glucose concentration of 12.4mmol/L. Over a mean CSII treatment duration of 14.4 days, glycemic control was generally favorable, with a median capillary blood glucose (CBG)derived time in range (3.9-10mmol/L, dTIR) of 84.1% (IQR, 75.0-90.7) and a median course-level mean glucose concentration of 7.1mmol/L (IQR, 6.4–8.0), while exposure to hypoglycemia remained low, with median derived time below range (dTBR) level 1 (3.0-3.9 mmol/L) of 1.7% (IQR, 0.0-3.8). The median prescribed total daily dose (TDD) of insulin was 0.49 U/kg/day on pump initiation, reached a course maximum of 0.73 U/kg/day, and decreased to 0.50 U/kg/day before CSII suspension. The broad ranges of baseline clinical characteristics, glycemic profiles, and insulin dosing reflect the clinical heterogeneity represented in the development dataset (Table 1).

**Table 1:** Clinical characteristics, glycemic profiles and insulin doses in the model-development cohort.

| Characteristics | Number of Cases | Mean±SD | Median (25%, 75%) | Range |
| --- | --- | --- | --- | --- |
| Age (year) | 1738 | 50.1±12.7 | 50.0 (41.0, 59.0) | 15.0–88.0 |
| Gender, male, N (%) | 1738 | 1181 (68.0%) | — | — |
| Body mass index (kg/m<sup>2</sup>) | 1734 | 24.9±3.5 | 24.6 (22.6, 26.8) | 15.4–43.0 |
| Waist circumference (cm) | 1392 | 90.3±9.7 | 90.0 (84.0, 96.0) | 61.0–140.0 |
| HbA1c (%) | 1708 | 10.9±2.3 | 10.9 (9.2, 12.6) | 5.5–20.0 |
| Fasting glucose (mmol/L) | 1579 | 12.4±3.9 | 12.1 (9.2, 14.8) | 5.5–30.0 |
| dTIR (3.9–10 mmol/L) (%) | 1737 | 81.1±13.3 | 84.1 (75.0, 90.7) | 6.7–100.0 |
| dTBR level 1 (3.0–3.9 mmol/L) (%) | 1737 | 2.6±3.1 | 1.7 (0.0, 3.8) | 0.0–24.8 |
| dTBR level 2 (<3.0 mmol/L) (%) | 1737 | 0.3±0.8 | 0.0 (0.0, 0.0) | 0.0–9.4 |
| Mean blood glucose (mmol/L) | 1737 | 7.4±1.4 | 7.1 (6.4, 8.0) | 4.9–18.0 |
| Mean fasting glucose (mmol/L) | 1737 | 6.0±1.0 | 5.8 (5.4, 6.4) | 3.9–15.4 |
| Mean 2h-postprandial glucose (mmol/L) | 1737 | 8.4±1.7 | 8.0 (7.2, 9.2) | 4.9–19.6 |
| Standard deviation of glucose (mmol/L) | 1737 | 2.6±0.9 | 2.5 (2.0, 3.1) | 0.7–6.5 |
| Coefficient of variance of glucose (%) | 1737 | 35.4±7.5 | 35.2 (30.3, 40.3) | 13.5–66.2 |
| Duration of CSII treatment (Days) | 1738 | 14.4±4.7 | 15.0 (11.0, 17.0) | 3.0–57.0 |
| Day 0 dose (U/kg) | 1729 | 0.50±0.15 | 0.49 (0.39, 0.59) | 0.04–1.53 |
| Maximum dose (U/kg) | 1729 | 0.77±0.21 | 0.73 (0.62, 0.88) | 0.18–2.00 |
| Suspension dose (U/kg) | 1729 | 0.52±0.23 | 0.50 (0.35, 0.65) | 0.05–1.54 |

Data are presented as mean ± SD, median (IQR), or n (%), as appropriate. Each CSII treatment course was counted as one case, including repeated courses from the same patient. Derived time in range and time below range were calculated from the 8-point capillary blood glucose measurements. Insulin doses represent calendar-day totals, with shorter infusion periods on pump-initiation and suspension days.

Across all treatment courses, 200216 scheduled CBG time points were identified. Of these, 148572 (74.2%) measurements were directly observed, and 13265 (6.6%) missing values were imputed using observations at the corresponding scheduled time point within the same treatment course. The remaining 38379 (19.2%) time points remained missing after this course-level preprocessing. The dataset also contained 35074 insulin pump prescription records, with a median of 17 records per course (IQR, 11-28; range, 2-97) and 18 per patient (IQR, 11-29; range, 2-101).

### Stage-specific model development and internal validation

Of the 1738 eligible treatment courses, 1239 (71.3%) had complete data for all five baseline clinical features (age, BMI, waist circumference, HbA1c, pre-CSII fasting glucose) and contributed to pump-initiation model development. Recorded initial basal rates and uniform meal-bolus doses were available for 1239 and 1231 instances, respectively. After stage-specific eligibility and temporal alignment were applied, sliding-window sampling generated 22970 one-day-history and 21292 two-day-history instances. Sample sizes differed slightly across prediction tasks according to the availability of the corresponding recorded doses. These datasets supported 18 independent LSTM models, comprising two models for pump initiation and 16 models for subsequent daily dose adjustment using one- or two-day glucose and insulin histories. Each history length supported eight models covering basal-rate and breakfast, lunch and dinner bolus recommendations through morning and afternoon decision pathways.

Internal validation using out-of-fold predictions showed clear differences across information stages. Compared with the corresponding pump-initiation tasks, history-based models had lower mean absolute error (MAE) and root mean squared error (RMSE), together with higher coefficients of determination (*R*^2^). At initiation, predictions explained more variation in basal rates than in uniform bolus doses. Once glucose and prescription histories were incorporated, agreement with recorded doses was consistently closer across both basal-rate and meal-bolus tasks. Performance was broadly similar between morning and afternoon pathways, although meal-specific differences remained. Breakfast-bolus errors were generally higher than lunch- and dinner-bolus errors in the one-day-history models. With two-day histories, errors were more uniform across meals, primarily reflecting lower breakfast-bolus errors rather than improvement in every task. Basal-rate errors also decreased with the additional history day, whereas lunch- and dinner-bolus performance showed smaller, inconsistent changes (Supplementary Table 1).

The prediction agreement plots further characterized these stage-specific differences. At pump initiation, predicted doses clustered toward the middle of the recorded dose range, particularly for uniform meal boluses, with a tendency to overestimate lower prescribed doses and underestimate higher ones. In contrast, history-based predictions were concentrated more closely around the line of identity across both decision pathways, indicating closer agreement with clinician-recorded doses. Nevertheless, this overall agreement was not uniform across individual records. Some predictions remained substantially above or below the recorded doses, and underprediction was still apparent among some of the higher recorded bolus doses. Thus, the closer aggregate agreement of the history-based models coexisted with discrepancies in individual recommendations (Supplementary Figure 1). Analysis by recorded dose-change category revealed additional differences not captured by overall prediction errors. Basal-rate models agreed more frequently with the direction of recorded decreases than increases, and two-day histories improved directional agreement in both categories. Bolus agreement depended on the meal and decision pathway. The additional history day improved breakfast-bolus agreement and afternoon lunch-bolus agreement, but did not produce a consistent improvement across all remaining tasks. Despite these differences in directional agreement, the mean dose differences followed a common pattern across all 16 history-based models. Predictions were higher than recorded prescriptions when clinicians reduced doses and lower when clinicians increased them, consistent with more moderate average adjustments relative to the preceding-day prescription. These discrepancies were smaller for two-day basal-rate, breakfast-bolus and afternoon lunch-bolus models than for their one-day counterparts, whereas changes in the remaining bolus tasks were less pronounced. For unchanged prescriptions, mean prediction differences were close to zero, yet predictions infrequently remained within the narrow numerical maintenance range. Small average differences therefore did not imply that individual prescriptions were preserved, distinguishing aggregate dose agreement from agreement in prescribing behavior (Fig. 2).

**Fig. 2.**
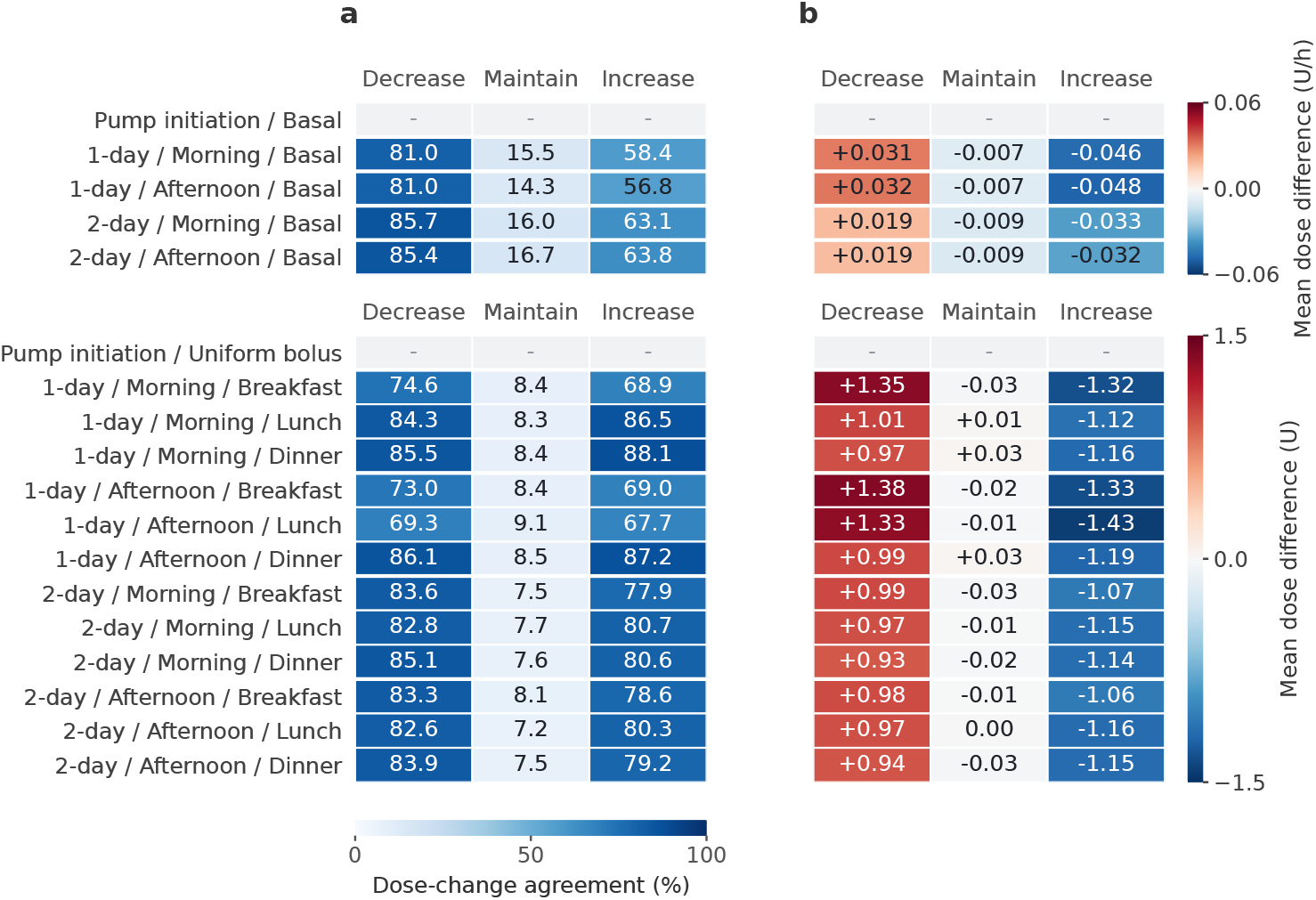
Internal validation of dose-adjustment direction and magnitude. Out-of-fold predictions are grouped by recorded dose decreases, maintenance and increases. (a) Agreement between predicted and recorded dose-change categories (%). (b) Mean predicted-minus-recorded dose. Changes were defined relative to the same preceding-day prescription, with maintenance defined as within

### Performance of model-directed treatment within the randomized clinical trial (RCT)

A total of 103 patients were randomized to the model-directed group. Of these, 29 discontinued the treatment before protocol completion, including six requiring physician takeover after meeting the prespecified hyperglycemia criterion (fasting glucose *>*10.0mmol/L on two consecutive days), 17 patients underwent earlier than scheduled CSII suspension at their request for early discharge. The remaining six patients were withdrawn from the study after conditions meeting the exclusion criteria were identified during hospitalization: one case each of latent autoimmune diabetes in adults (LADA), hyperthyroidism, cryptococcal infection, intestinal obstruction, pancreatitis, and coronary heart disease requiring transfer to another department. CSII was continued beyond the scheduled duration in 13 patients owing to ongoing treatment for comorbid conditions. After excluding the six patients who met the exclusion criteria, clinical outcomes were evaluated in the remaining 97, including those requiring physician takeover or receiving shorter or longer CSII courses than scheduled. Analyses were restricted to data collected during model-directed treatment. Among them, 36 had newly diagnosed and 61 had previously treated diabetes. Newly diagnosed patients were younger and had higher baseline HbA1c and eGFR, whereas anthropometric measures, fasting glucose, and random C-peptide before pump initiation did not differ significantly between groups. The overall median dTIR was 84.5%, with low proportions of sampled glucose values in the level 1 and level 2 hypoglycemic ranges. Newly diagnosed patients achieved higher dTIR and lower mean and 2-h postprandial glucose, together with lower glucose standard deviation (SD). However, fasting glucose during treatment, glucose coefficient of variation (CV), and hypoglycemic exposure did not differ significantly between groups. Insulin doses at initiation, at the within-course maximum, and at the end of model-directed treatment also showed no significant between-group differences (Table 2).

**Table 2:**
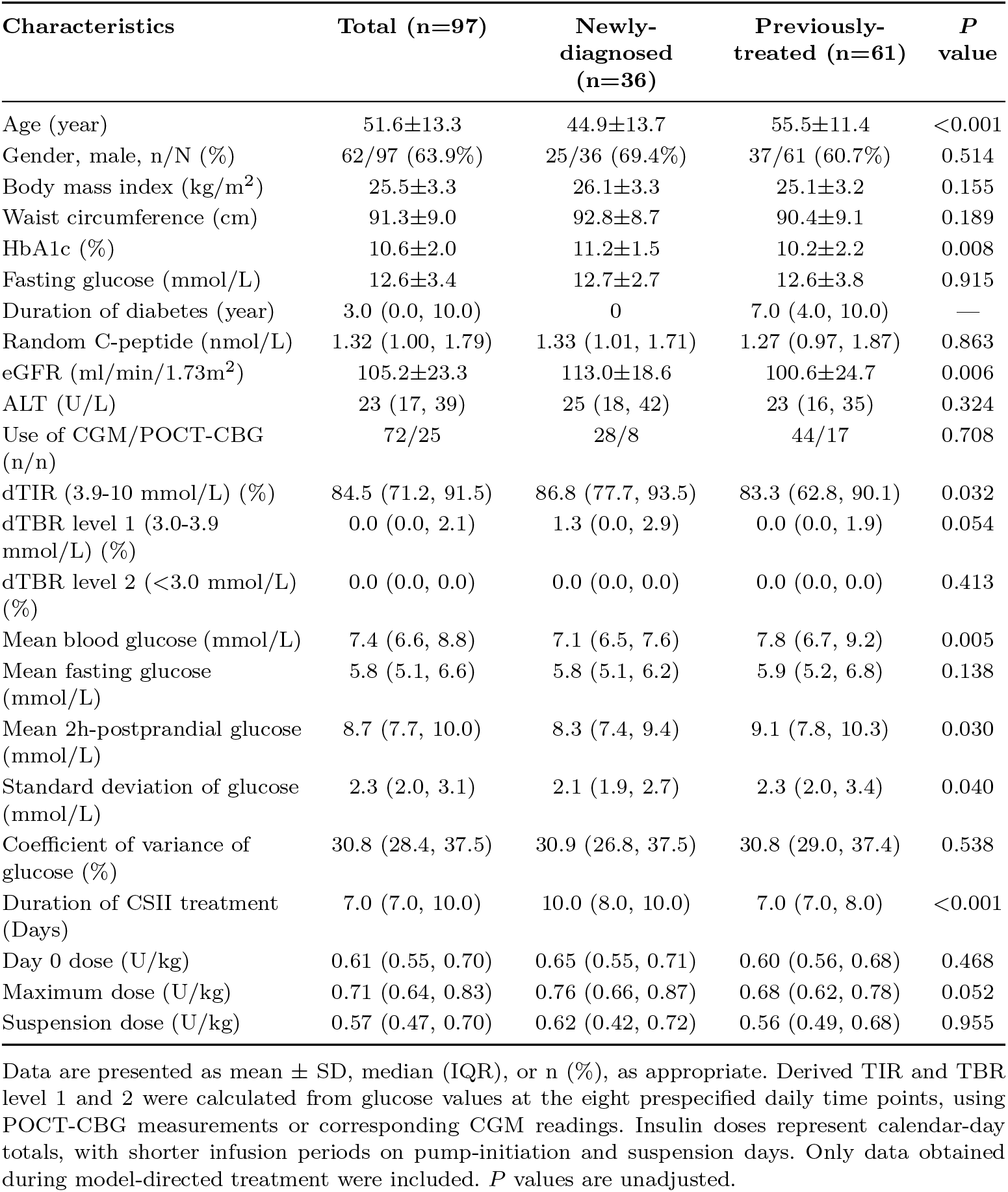
Clinical characteristics and performance of the model-directed group.

| Characteristics | Total (n=97) | Newly-diagnosed (n=36) | Previously-treated (n=61) | *P* value |
| --- | --- | --- | --- | --- |
| Age (year) | 51.6±13.3 | 44.9±13.7 | 55.5±11.4 | <0.001 |
| Gender, male, n/N (%) | 62/97 (63.9%) | 25/36 (69.4%) | 37/61 (60.7%) | 0.514 |
| Body mass index (kg/m<sup>2</sup>) | 25.5±3.3 | 26.1±3.3 | 25.1±3.2 | 0.155 |
| Waist circumference (cm) | 91.3±9.0 | 92.8±8.7 | 90.4±9.1 | 0.189 |
| HbA1c (%) | 10.6±2.0 | 11.2±1.5 | 10.2±2.2 | 0.008 |
| Fasting glucose (mmol/L) | 12.6±3.4 | 12.7±2.7 | 12.6±3.8 | 0.915 |
| Duration of diabetes (year) | 3.0 (0.0, 10.0) | 0 | 7.0 (4.0, 10.0) | — |
| Random C-peptide (nmol/L) | 1.32 (1.00, 1.79) | 1.33 (1.01, 1.71) | 1.27 (0.97, 1.87) | 0.863 |
| eGFR (ml/min/1.73m<sup>2</sup>) | 105.2±23.3 | 113.0±18.6 | 100.6±24.7 | 0.006 |
| ALT (U/L) | 23 (17, 39) | 25 (18, 42) | 23 (16, 35) | 0.324 |
| Use of CGM/POCT-CBG (n/n) | 72/25 | 28/8 | 44/17 | 0.708 |
| dTIR (3.9-10 mmol/L) (%) | 84.5 (71.2, 91.5) | 86.8 (77.7, 93.5) | 83.3 (62.8, 90.1) | 0.032 |
| dTBR level 1 (3.0-3.9 mmol/L) (%) | 0.0 (0.0, 2.1) | 1.3 (0.0, 2.9) | 0.0 (0.0, 1.9) | 0.054 |
| dTBR level 2 (<3.0 mmol/L) (%) | 0.0 (0.0, 0.0) | 0.0 (0.0, 0.0) | 0.0 (0.0, 0.0) | 0.413 |
| Mean blood glucose (mmol/L) | 7.4 (6.6, 8.8) | 7.1 (6.5, 7.6) | 7.8 (6.7, 9.2) | 0.005 |
| Mean fasting glucose (mmol/L) | 5.8 (5.1, 6.6) | 5.8 (5.1, 6.2) | 5.9 (5.2, 6.8) | 0.138 |
| Mean 2h-postprandial glucose (mmol/L) | 8.7 (7.7, 10.0) | 8.3 (7.4, 9.4) | 9.1 (7.8, 10.3) | 0.030 |
| Standard deviation of glucose (mmol/L) | 2.3 (2.0, 3.1) | 2.1 (1.9, 2.7) | 2.3 (2.0, 3.4) | 0.040 |
| Coefficient of variance of glucose (%) | 30.8 (28.4, 37.5) | 30.9 (26.8, 37.5) | 30.8 (29.0, 37.4) | 0.538 |
| Duration of CSII treatment (Days) | 7.0 (7.0, 10.0) | 10.0 (8.0, 10.0) | 7.0 (7.0, 8.0) | <0.001 |
| Day 0 dose (U/kg) | 0.61 (0.55, 0.70) | 0.65 (0.55, 0.71) | 0.60 (0.56, 0.68) | 0.468 |
| Maximum dose (U/kg) | 0.71 (0.64, 0.83) | 0.76 (0.66, 0.87) | 0.68 (0.62, 0.78) | 0.052 |
| Suspension dose (U/kg) | 0.57 (0.47, 0.70) | 0.62 (0.42, 0.72) | 0.56 (0.49, 0.68) | 0.955 |

Data are presented as mean ± SD, median (IQR), or n (%), as appropriate. Derived TIR and TBR level 1 and 2 were calculated from glucose values at the eight prespecified daily time points, using POCT-CBG measurements or corresponding CGM readings. Insulin doses represent calendar-day totals, with shorter infusion periods on pump-initiation and suspension days. Only data obtained during model-directed treatment were included. *P* values are unadjusted.

Among the 72 patients monitored with CGM, 28 had newly diagnosed and 44 had previously treated diabetes. Median TIR was high in both groups over the treatment course and exceeded 90% at pump suspension, with no significant between-group differences in TIR at initiation, over the treatment course, or at suspension. Newly diagnosed patients had lower mean glucose over the treatment course and at pump suspension, together with a lower glucose management indicator. Time above range and overall time below range did not differ significantly between groups, although level 1 TBR at pump suspension was higher in newly diagnosed patients. Level 2 hypoglycemic exposure remained low in both groups. Measures of glucose variability, including SD, CV, and mean amplitude of glycemic excursions, showed no significant between-group differences (Supplementary Table 2).

### Insulin dose and glucose trajectories

Insulin dose trajectories were examined alongside daily glucose profiles to characterize dose adjustment over the treatment course. In the historical development cohort, glucose declined rapidly after CSII initiation and then decreased more gradually, while insulin doses initially increased, plateaued, and subsequently declined. Newly diagnosed patients receiving model-directed treatment started at higher insulin doses than those in the historical development dataset and showed continued glycemic improvement, with modest early dose increases followed by a plateau and a more pronounced decline later in the observed course. In previously treated patients, the initial glucose reduction was followed by fluctuations at a higher level, while insulin doses remained relatively stable with modest upward adjustments. Although median day-to-day dose changes were generally small, the historical and newly diagnosed groups showed an overall decline in insulin dose across the later observation period (Fig. 3). Individual-level analysis revealed substantial dose reductions in some previously treated patients despite the absence of a clear decline in the group-level trajectory. Earlier attainment of the maximum insulin dose was associated with greater subsequent dose reduction in all three groups (all *P <* 0.001). Longer treatment duration was also associated with greater dose reduction in the historical cohort and previously treated patients, but no significant association was observed in newly diagnosed patients (Supplementary Figure 2).

**Fig. 3.**
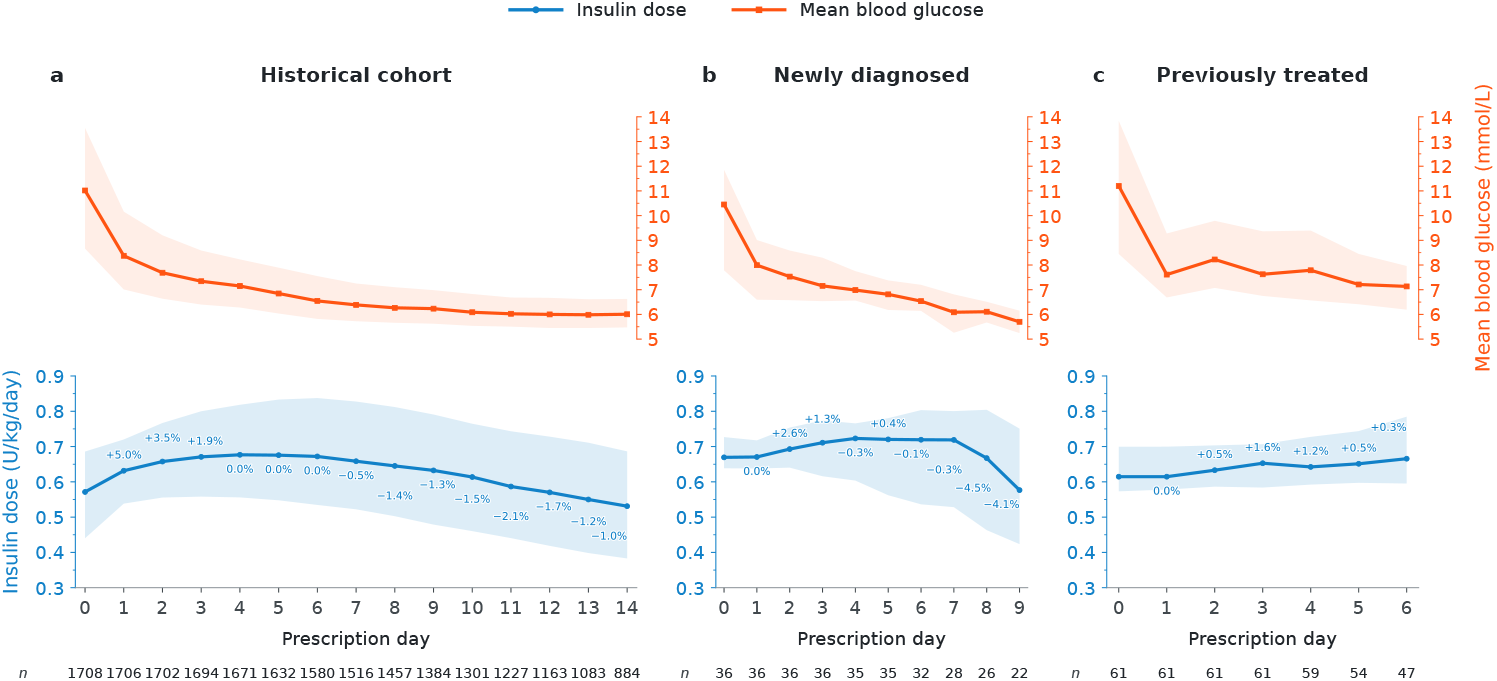
Glucose and insulin dose trajectories during CSII. (a) Historical development cohort. (b) Newly diagnosed and (c) previously treated patients in the model-directed group. Lines and shaded bands represent medians and interquartile ranges of total daily insulindose (blue) and individual daily mean glucose (orange). Insulin doses were calculated from each prescription over a full 24 h, including on pump-initiation and discontinuation days. Glucose values represent each patient’s mean glucose on the corresponding calendar day. Percentages indicate median within-patient dose changes from the preceding prescription day and may therefore differ in direction from changes in the plotted daily medians. Numbers below the panels indicate treatment courses in (a) and patients in (b, c) with both glucose and insulin dose data available.

### Development-data representativeness and early glycemic response according to achieved dTIR

To explore factors associated with suboptimal glycemic response, clinical characteristics and treatment profiles were compared between patients with dTIR *≥* 70% (n=73) and *<* 70% (n=24). Newly diagnosed diabetes was more common in the higher-dTIR group, which was also characterized by younger age, shorter diabetes duration, higher BMI and waist circumference, lower baseline fasting glucose, and higher eGFR. Baseline HbA1c and random C-peptide did not differ significantly between groups. Patients with dTIR *<* 70% had higher mean, fasting, and postprandial glucose and greater glucose SD, whereas differences in glucose coefficient of variation and hypoglycemic exposure were not statistically significant. Initial insulin doses did not differ significantly, but maximum doses and doses at the end of model-directed treatment were higher in the lower-dTIR group (Table 3).

**Table 3:** Clinical characteristics, glycemic profiles, and insulin doses of patients under model-directed treatment by dTIR category.

| Characteristics | dTIR $\geq 70\%$ | dTIR $< 70\%$ | *P* value |
| --- | --- | --- | --- |
| Number of patients, N | 73 | 24 |  |
| Newly diagnosed/Previously treated (N/N) | 33/40 | 3/21 | 0.004 |
| Age (year) | 48.4±13.2 | 61.2±7.9 | <0.001 |
| Gender, male, N (%) | 49 (67.1%) | 13 (54.2%) | 0.252 |
| Body mass index (kg/m<sup>2</sup>) | 26.2±2.9 | 23.3±3.4 | <0.001 |
| Waist circumference (cm) | 92.7±7.9 | 87.0±10.8 | 0.024 |
| HbA1c (%) | 10.4±2.0 | 11.0±1.9 | 0.191 |
| Fasting glucose (mmol/L) | 12.2±3.3 | 14.0±3.5 | 0.028 |
| Duration of diabetes (year) | 0.5 (0.0, 5.0) | 9.5 (4.8, 13.0) | <0.001 |
| Random C-peptide (nmol/L) | 1.33 (1.01, 1.77) | 1.28 (0.80, 1.81) | 0.662 |
| eGFR (ml/min/1.73m<sup>2</sup>) | 109.2±21.8 | 93.3±24.0 | 0.007 |
| ALT (U/L) | 23 (16, 39) | 24 (19, 34) | 0.880 |
| Use of CGM/POCT-CBG (N/N) | 54/19 | 18/6 | 0.920 |
| Derived TIR (3.9-10 mmol/L) (%) | 87.2 (83.1, 93.6) | 58.1 (50.0, 63.8) | <0.001 |
| Derived TBR level 1 (3.0-3.9 mmol/L) (%) | 0.0 (0.0, 2.2) | 0.0 (0.0, 2.1) | 0.194 |
| Derived TBR level 2 (<3.0 mmol/L) (%) | 0.0 (0.0, 0.0) | 0.0 (0.0, 0.0) | 0.194 |
| Mean blood glucose (mmol/L) | 7.0 (6.5, 7.6) | 9.4 (9.0, 10.2) | <0.001 |
| Mean fasting glucose (mmol/L) | 5.6 (5.0, 6.1) | 6.8 (6.4, 8.0) | <0.001 |
| Mean 2h-postprandial glucose (mmol/L) | 8.0 (7.4, 9.0) | 10.7 (10.0, 12.1) | <0.001 |
| Standard deviation of glucose (mmol/L) | 2.1 (1.9, 2.5) | 3.4 (2.9, 3.8) | <0.001 |
| Coefficient of variation of glucose (%) | 30.5 (28.2, 34.7) | 34.0 (30.5, 40.3) | 0.057 |
| Day 0 dose (U/kg/d) | 0.61 (0.54, 0.69) | 0.65 (0.59, 0.72) | 0.301 |
| Maximum dose (U/kg/d) | 0.70 (0.62, 0.77) | 0.80 (0.67, 0.94) | 0.019 |
| Suspension dose (U/kg/d) | 0.55 (0.43, 0.65) | 0.74 (0.63, 0.84) | <0.001 |

Data are presented as mean ± SD, median (IQR), or n (%), as appropriate. *P* values are unadjusted.

To examine whether suboptimal control was apparent early in treatment, mean glucose during the first 24 h was assessed in relation to dTIR over the subsequent 24-72-h window, alongside insulin dose trajectories over the first 72 h. Higher first-24-h mean glucose was associated with lower dTIR over the subsequent 24-72 h (Spearman’s *ρ* = *−*0.567, *P <* 0.001). Patients with whole-course dTIR *<* 70% had higher glucose from the first 24 h onward, with differences persisting across all three windows (all Holm-adjusted *P <* 0.001). Despite this early glycemic separation, initial total insulin doses were similar between groups. By the third 24-h window, median basal and bolus doses had increased in the lower-dTIR group, while doses in the higher-dTIR group remained relatively stable. Between-group differences in insulin doses were not statistically significant during these early windows (Fig. 4), although maximum and end-of-treatment doses over the full observed course were higher in the lower-dTIR group (Table 3).

**Fig. 4.**
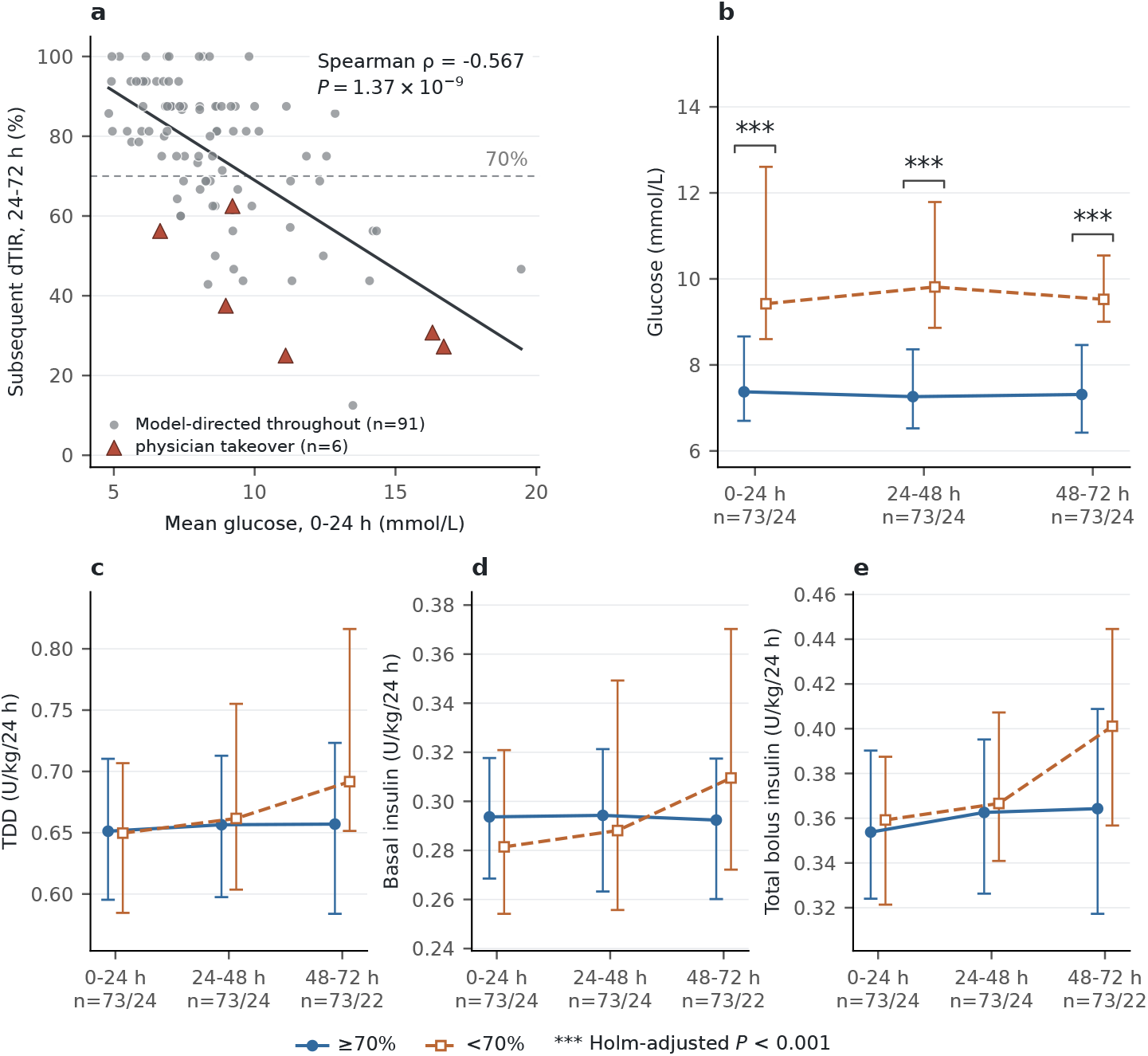
Early glycemic response and insulin dose trajectories. (a) First-24-h mean glucose in relation to subsequent 24–72-h dTIR. (b–e) Glucose, total daily insulin dose (TDD), basal insulin, and total bolus insulin across three consecutive 24-h windows, grouped by whole-course dTIR (*≥* 70% vs. *<* 70%). Points and error bars in b–e represent medians and interquartile ranges; triple asterisks indicate Holm-adjusted *P <* 0.001. Triangles in panel a identify patients requiring physician takeover for persistent hyperglycemia; the solid line indicates a linear fit. Only model-directed treatment data were included. Two incomplete final dose windows resulting from takeover were excluded from dose summaries.

The k-nearest-neighbor (kNN) distance was used to assess patients’ similarity to the development data, with larger distances indicating lower local representation. Based on baseline clinical features alone, this distance showed only a weak inverse association with subsequent 24–72-h dTIR (*ρ* = *−*0.200, Holm-adjusted *P* = 0.0495). Patients with markedly different glycemic outcomes had overlapping distances, and those requiring physician takeover were not confined to the most distant observations. When first-24-h glucose data were incorporated, the inverse association was more pronounced (*ρ* = *−*0.420; Holm-adjusted *P <* 0.001): larger distances generally corresponded to poorer subsequent glycemic control, whereas higher dTIR values were more common at smaller distances. Nevertheless, some patients had low dTIR despite relatively small distances, indicating that proximity to the development data did not consistently distinguish patients with favorable and unfavorable glycemic responses (Fig. 5).

**Fig. 5.**
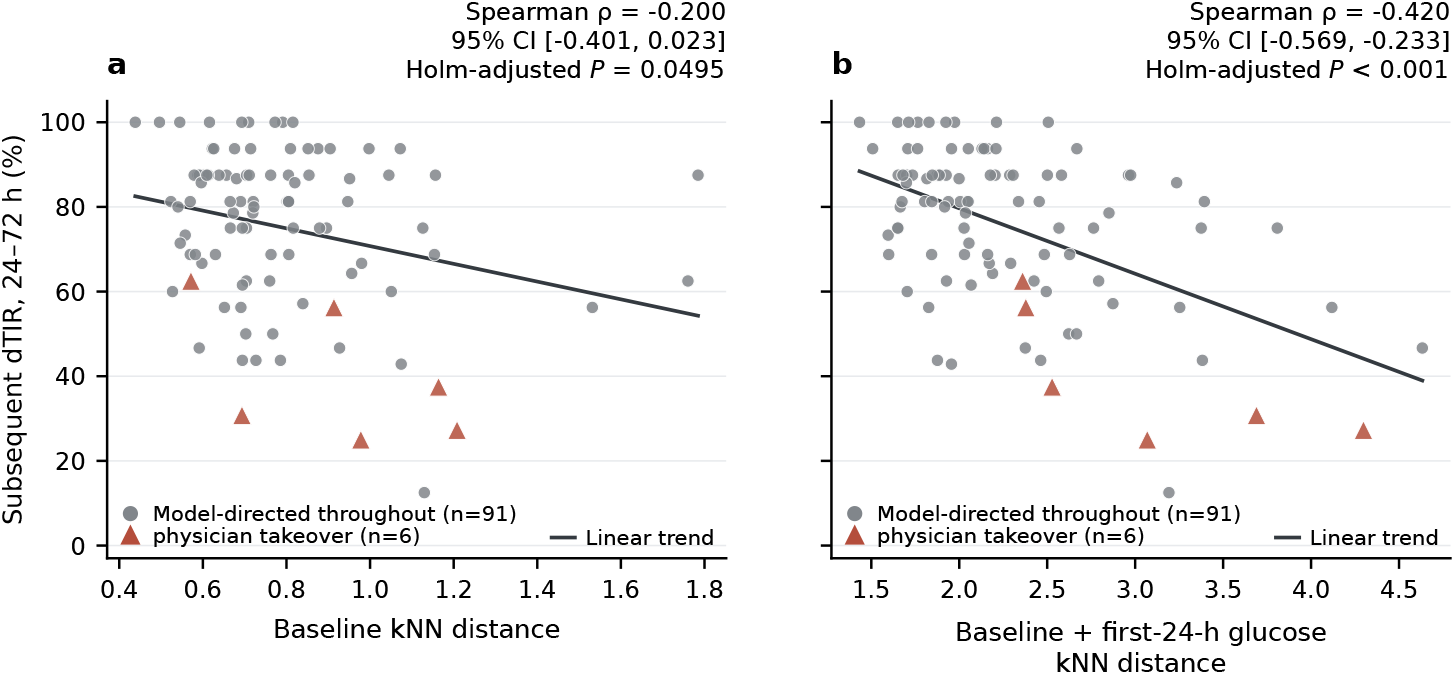
Development-data representativeness and subsequent glycemic control. Associations between subsequent 24–72-h dTIR and k-nearest-neighbor (kNN) distances calculated using baseline clinical features alone (a) or combined with first-24-h glucose data (b). Larger distances indicate lower local representation in the development data. Triangles identify patients requiring physician takeover for persistent hyperglycemia; circles represent patients remaining under model-directed treatment. Only model-directed treatment data were included. Solid lines indicate linear fits.

## 3 Discussion

This study demonstrates how accumulated clinical expertise can be translated into a data-driven system that guides insulin dosing throughout open-loop CSII and achieves excellent glycemic control. Developed from more than two decades of clinical records, the LSTM system generated initial prescriptions from baseline patient characteristics and directed subsequent titration using evolving glucose and insulin histories. Internal validation showed close agreement with recorded prescriptions for history-based adjustments, while prospective clinical application achieved high time in range with limited hypoglycemic exposure. Favorable outcomes extended beyond newly diagnosed diabetes to patients with longer-standing, previously treated disease, supporting the clinical value of the approach across differing treatment histories. Beyond overall performance, early glucose responses distinguished patients with subsequent suboptimal control, and their incorporation into development-data distance strengthened its association with subsequent glycemic outcomes. Together, these findings establish a clinically grounded foundation for model-directed insulin titration and identify opportunities to refine its use through early recognition of patients requiring additional support.

The value of a data-driven dosing system depends not only on its architecture but also on the quality and clinical relevance of its development data. The present dataset grew out of more than two decades of sustained clinical practice and research in CSII-based short-term IIT at the study center, spanning early investigations of drug-free glycemic control and *β*-cell recovery through to multicenter evaluation of an intensive treatment strategy followed by simplified maintenance therapy [12, 13]. It linked patient characteristics to repeated glucose measurements and insulin prescriptions, capturing dose escalation during correction of hyperglycemia and subsequent reduction as control improved. The favorable glycemic outcomes in the historical cohort provide a clinically meaningful foundation for learning these prescribing patterns. Combining clinical research with routine practice also captured varied patient characteristics and treatment responses within the center’s population. Together, this longitudinal detail and accumulated experience supported the development of context-dependent dosing recommendations. The contribution lies in translating an established body of specialist practice into a reproducible treatment approach that could support clinicians with less experience in pump-based insulin therapy.

Internal validation showed that estimation at pump initiation was more challenging than subsequent titration informed by recent glucose and insulin histories. Initial recommendations relied on five baseline characteristics, whereas subsequent adjustments were informed by recent glucose profiles and the preceding insulin regimen. This additional context provides a clinically plausible explanation for the closer agreement of history-based predictions with recorded prescriptions. Beyond overall agreement, the direction-specific analyses revealed a tendency toward more moderate adjustments, with both dose increases and reductions attenuated relative to clinician-recorded changes. Such behavior may favor gradual titration, but could also delay correction when a larger increase is needed to address persistent hyperglycemia. Smaller individual reductions do not preclude substantial cumulative dose reductions over successive treatment days. The clinical implications therefore depend on how these adjustments translate into glucose control over time, rather than on prescription agreement alone. Agreement with historical prescriptions reflects consistency with clinical dosing practice, whereas prospective glycemic outcomes provide evidence of the model’s clinical utility.

The clinical contribution of this system lies in bringing both insulin initiation and subsequent titration under a unified model-directed workflow. In the iNCDSS randomized trial, insulin regimens were initiated or modified by endocrinologists during a one-day run-in before algorithm-guided titration, which subsequently achieved glycemic control noninferior to senior physician care [7, 14]. Similarly, the RL-DITR proof-of-concept trial introduced model-guided titration after the first treatment cycle [15]. Another randomized trial evaluated learning-based meal bolus recommendations, but starting doses and ongoing basal insulin adjustments remained physician-directed [16]. A retrospective study used routine electronic health records to estimate initial inpatient insulin requirements more accurately than conventional weight-based formulas, although its clinical utility remained to be prospectively evaluated [17]. A conference abstract also reported externally validated machine learning models for initial insulin pump dosing[18].Our study developed and prospectively evaluated a dosing system that integrates initial prescribing with subsequent titration for open-loop CSII. It generates the initial pump prescription from five routinely available clinical characteristics and subsequently recommends daily basal-rate and premeal bolus adjustments using updated glucose and insulin histories. The system provides a consistent basis for dosing from the first prescription onward, reducing reliance on individual prescribing experience while preserving physician-led safety monitoring and intervention when needed. This is particularly relevant to the wider implementation of model-directed insulin pump therapy in settings with limited specialist expertise.

The glucose–insulin trajectories illustrate two complementary demands of short-term IIT, sufficient early dosing to correct hyperglycemia and timely dose reduction as control improves. Previous CSII research linked the starting dose to time to glycemic control, with faster control associated with more hypoglycemia [19]. Both model-directed subgroups received higher initial doses than the historical cohort, followed by smaller early increments. This pattern is consistent with the center’s evolving practice of using higher starting doses to facilitate earlier control and allow more time for subsequent dose reduction. Smaller increments therefore do not necessarily indicate conservative treatment, although escalation may still have been insufficient for some patients with persistent hyperglycemia. Later improvements in insulin sensitivity and *β*-cell function provide a physiological basis for declining insulin requirements [2, 20]. Previous studies have linked dose reductions after glycemic normalization to improved insulin resistance and sustained remission [21], documented a transition from escalation to reduction [22], and highlighted the need for timely down-titration when hypoglycemia occurs despite ongoing reductions [23]. Our individual-level analyses identified reductions in both clinical subgroups, with larger reductions associated with earlier attainment of the maximum dose. In previously treated patients, greater individual dose reductions were associated with longer treatment, suggesting that their shorter scheduled courses may partly explain the lack of a clear decline in the group-level dose trajectory. In newly diagnosed patients, greater reductions were associated with earlier dose peaks, whereas the association with treatment duration was less evident within the observed treatment window. Together, these findings support glucose-responsive adjustment in both directions, while highlighting the opportunity for more timely escalation in patients whose glucose remains elevated.

Practical flexibility also extends to the source of glucose inputs. Although developed using CBG measurements, the system was subsequently applied with either CBG or CGM-derived glucose values at corresponding time points. This experience supports the feasibility of model-directed dosing across different monitoring settings, although the two input sources were not directly compared.

Although the development data were drawn largely from newly diagnosed patients, approximately two-thirds of previously treated patients achieved a dTIR *≥* 70%, supporting the system’s potential applicability beyond newly diagnosed diabetes. Patients with suboptimal control were older and leaner, had longer diabetes duration and presented with higher baseline glucose levels. These findings broadly align with previous observations linking clinical characteristics to the response to short-term IIT and subsequent remission [3, 24–27]. Such characteristics may help identify patients who warrant closer monitoring, but do not by themselves define the limits of model-directed treatment.

Early glycemic response provided additional information beyond these baseline characteristics. Higher mean glucose during the first 24 h was associated with poorer control over the subsequent 24–72 h, suggesting that an inadequate response could become apparent well before completion of treatment. Similar random C-peptide concentrations between groups do not necessarily imply equivalent *β*-cell compensation, because C-peptide must be interpreted in relation to the glucose level at the time of measurement and the degree of insulin resistance [28]. Early glucose profiles reflect the combined effects of endogenous insulin secretion, insulin sensitivity and administered insulin, and may therefore reveal a mismatch between the current regimen and metabolic needs that a single C-peptide measurement does not fully capture. Persistent early hyperglycemia could thus provide a practical signal for timely clinical reassessment, although specific intervention thresholds require prospective evaluation.

The kNN analysis explored whether suboptimal glycemic control was associated with limited representation in the development data. A large dataset may still provide limited coverage of particular patient profiles. Studies using electronic health records have shown that targeted expansion or augmentation of training data can improve performance in underrepresented or poorly performing subgroups, highlighting the importance of subgroup coverage beyond overall sample size [29, 30]. In the present study, baseline distance was only weakly associated with subsequent dTIR, indicating that patients whose baseline profiles were less similar to historical cases did not consistently experience poorer control. Although the development data predominantly represented newly diagnosed diabetes, previously treated patients may share similar glucose profiles and insulin requirements. Clinical differences between response groups therefore do not necessarily imply that the characteristics captured by the model are poorly represented in its development data. When first-24-h glucose measurements were included, greater distance showed a numerically stronger association with poorer subsequent control. Similarity to historical cases was more closely associated with subsequent glycemic control when assessed using both baseline characteristics and first-24-h glucose profiles. However, the stronger association may partly reflect the prognostic information carried by early glucose itself, and the incremental value of the distance measure remains uncertain. These findings suggest that the applicability of the system should be assessed using both baseline characteristics and early treatment responses. This approach aligns with calls to identify inputs that differ from model-development data as a safeguard for clinical AI use [31]. Our findings provide a basis for incorporating early glucose responses into the assessment of system applicability and for developing criteria to guide timely clinical review. Such criteria will require prospective validation before routine use.

We developed this system to make experience-based CSII titration more accessible by supporting both the initial prescription and subsequent daily adjustments.Our approach places clinicians primarily in a supervisory role, focusing on safety monitoring and intervention when clinically indicated rather than routine dose adjustment. System-generated recommendations are followed without routine physician modification. This differs from the AI-DSS evaluated by Nimri et al. [32], in which algorithm-generated pump adjustments were reviewed and approved by healthcare professionals before being communicated to patients. The d-Nav system similarly combined automated titration of injection-based insulin therapy with professional support in type 2 diabetes [6, 33]. We brought this principle to short-term inpatient CSII, where glucose levels and insulin doses can change rapidly. The small number of patients requiring takeover for persistent hyperglycemia and the low frequency of hypoglycemia support the feasibility of this approach and its potential to reduce the burden of daily dose determination. Our early-response findings suggest a collaborative strategy for future evaluation. For patients with persistent early hyperglycemia, physicians could temporarily direct dose adjustment, with their prescriptions and corresponding glucose measurements entered into the system as updated treatment histories. Once control improves, model-directed dosing could resume using these recent records. Prospective evaluation could establish when to intervene and when to resume model-directed treatment, allowing clinical expertise to complement model support when most needed.

Several limitations should be acknowledged. First, model development and prospective clinical evaluation were conducted at a single center, using development data predominantly derived from newly diagnosed patients. Although the clinical findings support potential use in previously treated patients, performance across institutions with different patient populations and prescribing practices remains to be established. Experience with other clinical prediction systems underscores the importance of independent external validation and monitoring for changes in data distributions after deployment [34, 35]. Second, training on clinician-recorded prescriptions allowed the model to learn from accumulated dosing experience, but did not directly optimize subsequent glycemic outcomes. Prospective evaluation assessed the clinical effectiveness of this approach, although it did not establish whether the recommended doses were optimal. Third, we used a fixed training configuration without systematic hyperparameter optimization or comparison with alternative architectures. The current implementation also recommends a single basal rate rather than a time-varying basal profile, limiting its ability to accommodate within-day differences in insulin delivery needs. Fourth, the different scheduled treatment durations in newly diagnosed and previously treated patients may partly account for differences in glycemic outcomes between these groups. Comorbidities and concomitant therapies may also have influenced the observed outcomes. Finally, the early-response and kNN analyses were exploratory and based on a modest clinical sample. They provide clinically relevant clues to suboptimal response but do not establish causal mechanisms or validated thresholds for intervention.

In conclusion, we translated accumulated clinical experience into an LSTM-based system that supports insulin dosing from CSII initiation through daily titration. Prospective use demonstrated effective glycemic control in both newly diagnosed and previously treated patients, while early glucose responses helped identify those who may require closer clinical oversight. These findings support the potential of model-directed CSII to standardize insulin pump therapy in patients with type 2 diabetes and reduce reliance on experienced clinicians for routine dose adjustment. Building on this foundation, alternative algorithms and time-varying basal profiles should be evaluated against the current system to improve early titration while preserving safety and timely dose reduction. Extending intensive insulin therapy through supervised outpatient continuation warrants investigation to determine whether longer treatment increases the likelihood of drug-free remission or simplifies subsequent treatment. Building on the learned relationship between glucose and insulin requirements, future work could explore real-time dosing using continuous CGM data as a step toward closed-loop insulin delivery. Multicenter studies, including hospitals with limited CSII experience, will be important to establish generalizability and quantify the benefits for clinical workflow.

## 4 Methods

### Data source and model-development cohort

Data of inpatients with newly diagnosed type 2 diabetes who received CSII-based short-term IIT at the Department of Endocrinology, the First Affiliated Hospital of Sun Yat-sen University between September 2001 and December 2022 were collected, including clinical characteristics, routine eight-point CBG measurements during CSII treatment, and daily insulin pump prescriptions comprising basal and premeal bolus doses. CBG was measured before and 2 hours after each meal, at bedtime, and at 3:00 a.m.; random measurements obtained at other times were excluded. The dTIR was calculated as the percentage of measurements within 3.9-10.0mmol/L divided by the sum of CBG counts [36, 37]. Likewise, the dTBR 1 (3.0-3.9mmol/L) and dTBR 2 (*<*3.0mmol/L) quantified level 1 and level 2 hypoglycemia frequencies. Routine CBG monitoring transitioned from four-point profiles (fasting and three 2-h postprandial measurements) before 2008 to eight-point profiles from 2008 onward, which comprised most of the development data. For retrospective model development, missing glucose values were handled separately at each scheduled time point within each treatment course. For time points other than fasting and 03:00, gaps bounded by observed values were linearly interpolated across treatment days, including consecutive gaps; leading gaps were filled using the first subsequent observed value at the same time point. Trailing gaps and time points with no observations throughout a course remained missing, as did missing fasting and 03:00 measurements at this preprocessing stage. This approach was intended to improve the completeness of historical glucose profiles while preserving their temporal structure and limiting imputation-related bias. This course-level procedure could use observations from later treatment days. The TDD on the day of CSII initiation was calculated and termed ‘Day 0 dose (U/kg/d)’. Similarly, the highest and final TDD per kilogram was termed ‘Maximum dose (U/kg/d)’ and ‘Suspension dose (U/kg/d)’. Some patients underwent repeated courses of short-term IIT when glycemia worsened after prolonged periods without glucose-lowering medication. Because their age, BMI, waist circumference, glycemic status had changed between admissions, each admission was treated as a distinct treatment course. However, all courses belonging to the same patient were linked using a unique patient identifier and assigned to the same training, validation, or test partition and cross-validation fold to prevent patient-level data leakage.

### Stage-specific LSTM model development

Stage-specific samples were constructed from each aligned treatment course. Within each calendar day, hourly basal-rate records were summarized as a single daily basal rate (U/h) by averaging the observed hourly settings, including explicitly recorded zeros, and rounding to two decimal places. For prescriptions with multiple basal-rate segments, this represented a duration-weighted mean over the recorded hours. Missing hours were excluded from the calculation, and days with no recorded settings had an undefined daily basal rate. The resulting daily value was used as the basal-rate prediction target and, for history-based models, as the corresponding historical basal-rate input. Pump-initiation inputs comprised five baseline features, including age, BMI, waist circumference, HbA1c and pre-CSII fasting glucose. After pump initiation, morning and afternoon decision pathways accommodated differences in admission timing and the availability of sufficient glucose data. The morning pathway used eight glucose measurements from pre-lunch on the preceding day to 2-h post-breakfast on the current day, whereas the afternoon pathway extended from pre-dinner on the preceding day to 2-h post-lunch on the current day. Each history step combined these glucose values with the basal rate and three meal boluses recorded on the preceding calendar day and the five baseline features, yielding 17 input features.

Sliding-window sampling generated one-day histories containing the latest input step and two-day histories containing two consecutive steps in chronological order. Windows advanced by one day, with sampling restricted to the first 20 target days per course and history length to limit the contribution of longer courses. The corresponding clinician-recorded dose on the target day served as the prediction target. Historical insulin inputs were taken from recorded prescriptions rather than model-generated doses. Samples were assessed for eligibility separately for each prediction task. Missing target doses were not imputed, whereas explicitly recorded zero doses were retained.

The framework comprised 18 independent regression models. Two pump-initiation models predicted the basal rate and a uniform bolus dose for the three meals. At each history-based stage, separate models predicted the basal rate and breakfast, lunch and dinner boluses for each decision pathway, yielding eight models per stage. Clinically, the one-day-history models supported the first post-initiation adjustment, although their training samples were drawn from eligible decision points throughout treatment. Once two history steps were available, the two-day-history models were applied on a rolling basis using the two most recent steps. The four predictions from the selected pathway formed the recommended basal–bolus prescription.

Each model comprised a single LSTM layer with 50 hidden units and a linear scalar output. Input dimensions were 1*×*5 for pump initiation, 1*×*17 for one-day histories and 2 *×* 17 for two-day histories. Numeric features were standardized separately for each task using parameters estimated from the training partition of each cross-validation fold. History-lag features were standardized separately, and remaining missing history-based inputs were replaced with the corresponding training-feature means. Pump-initiation models required complete data for all five baseline features. All models were trained for 200 epochs using mean squared error loss and Adam, with a learning rate of 0.001 and a batch size of 200. Training records were shuffled between epochs. The same configuration and model seed of 42 were used across tasks, with final-epoch weights retained without early stopping or validation-based epoch selection. Model fitting used PyTorch, and data processing and metric calculations used NumPy, pandas and scikit-learn.

### Internal validation

Patient-grouped five-fold cross-validation used a single fold assignment shared across all tasks, with seed 42. All courses and windows from each patient remained in the same fold. In each iteration, feature standardization, residual mean imputation and model fitting used four folds, and predictions were generated for the remaining fold. Five models were fitted per task, yielding 90 cross-validation models and one out-of-fold prediction for every eligible record. No best fold or seed was selected. After cross-validation, preprocessing parameters and one final model per task were refitted using all eligible development records, yielding 18 models for clinical deployment. These final models were not used to estimate validation performance.

Performance was assessed exclusively using pooled out-of-fold predictions within each task, with clinician-recorded prescriptions as the reference. MAE, RMSE and *R*^2^ were calculated from unrounded predictions, assigning equal weight to each eligible record. Percentile 95% confidence intervals were obtained from 2000 patient-level cluster bootstrap replicates, retaining all associated courses and records whenever a patient was sampled. Bootstrap analyses used a base seed of 123 with fixed task-specific offsets. Intervals were conditional on the fixed out-of-fold predictions and did not incorporate model-refitting variability. Prediction agreement across the recorded dose range was visualized using hexagonal-bin plots against the line of identity.

Dose-adjustment behavior was evaluated separately for recorded decreases, maintenance and increases. Both recorded and predicted changes were calculated relative to the same preceding-day dose of the corresponding insulin component. Maintenance was defined as a change within *±*0.005 U/h for basal rate or *±*0.05 U for bolus doses, corresponding to half the recording resolution. Changes below or above these ranges were classified as decreases or increases, respectively. These numerical thresholds did not represent clinical equivalence or safety limits. Within each recorded-change category, directional agreement was the percentage of predictions assigned to the same category, and mean dose difference was the average predicted-minus-recorded dose. Pump-initiation models were not assessed because no preceding-dose reference was available.

### Randomized clinical evaluation of model-directed insulin dosing

Following internal validation, the LSTM-based system was prospectively evaluated in a 24-patient feasibility study. The findings informed further model refinement and the eligibility and safety procedures for the subsequent RCT. These patients were included in model development but excluded from the RCT. The final system was fixed before RCT enrollment and remained unchanged throughout the trial. This prospective, single-center, parallel-group RCT compared model-directed with physician-directed insulin dosing during CSII treatment. Patients were stratified into newly diagnosed and previously treated diabetes with inadequate glycemic control groups, and randomly assigned 1:1 within each stratum using computer-generated permuted blocks of four. CSII was initiated on admission and scheduled for 9 days in newly diagnosed patients and 6 days in previously treated patients. Within hospital length-of-stay constraints, the longer course in newly diagnosed patients was intended to allow more time for glycemic normalization and facilitate subsequent drug-free remission. CSII was suspended after dinner bolus, and patients were discharged the following day. The study was approved by the ethics board of the hospital ([2024]-836), and all participants provided written informed consent. The trial was registered at ClinicalTrials.gov (NCT07066891).

In the model-directed group, basal and bolus doses throughout treatment were prescribed according to the system’s recommendations, without dietary caloric calculations or additional discretionary adjustments by physicians. Physicians provided diabetes education and safety monitoring while adhering to model-recommended doses, intervening only when severe hypoglycemia was anticipated, and taking over dose adjustment if prespecified hyperglycemia criteria required withdrawal from model-directed treatment. In the physician-directed group, doses were determined by the treating physician under senior supervision, without restrictions on segmented basal rates, supplemental insulin, or other clinically indicated adjustments. The majority of the patients used real-time CGM; those who declined CGM underwent routine eight-point CBG monitoring, with the corresponding eight daily glucose values used as model inputs. CGM data were available in real time to clinicians and patients in both groups. Full RCT outcomes will be reported separately, the present study focuses on system performance and exploration of its clinical applicability boundaries.

### Insulin dose and glucose trajectories

A post hoc descriptive analysis examined insulin dose and glucose trajectories in the historical development cohort and during model-directed treatment, with clinical patients stratified as newly diagnosed or previously treated. Daily total insulin doses (U/kg/day) were summarized by prescription day and paired with mean glucose values from the corresponding calendar day; the two measures did not necessarily cover identical 24-h intervals. Within each treatment course, percentage changes in insulin dose between consecutive prescription days were calculated and summarized as medians at each time point. Summaries used available observations, with the number of contributing courses (in historical development cohort) or patients (in RCT) reported for each day. An exploratory analysis examined individual-level insulin dose reduction, calculated as the percentage decrease from the maximum prescribed daily dose to that on the day of pump discontinuation. Both doses were standardized to 24 h to avoid apparent reductions attributable to the shorter infusion duration on the final day. Associations with time to first attainment of the maximum dose (Tmax) and duration of pump treatment were assessed separately for historical treatment courses and newly diagnosed and previously treated patients. Clinical analyses were restricted to the period of model-directed treatment, excluding observations after physician takeover.

### Assessment of early glycemic response and development-data representativeness

To explore factors associated with suboptimal glycemic control, patients were grouped by dTIR (*≥* 70% vs *<* 70%) over the observed course of model-directed treatment. Baseline clinical characteristics, treatment glucose profiles, and insulin doses were compared between groups. Early glycemic response was then assessed by relating mean blood glucose during the first 24 h after CSII initiation to dTIR over the subsequent 24–72 h. Glucose profiles and weight-adjusted total, basal, and bolus insulin doses were summarized across three consecutive 24-h windows within each whole-course dTIR group. Only data obtained while model-directed treatment remained active were included; incomplete dose windows resulting from physician takeover were excluded from 24-h dose summaries.

Representativeness was explored using kNN distances from each patient to the historical development data, considering the five baseline clinical features alone or combined with eight first-24-h glucose values. For each historical patient, the first successfully parsed course with recorded basal-rate data, ordered by course identifier, was selected. Patients were included in the reference set only if the selected course had complete data for all five baseline features and observed glucose values at at least six of the eight scheduled time points within the first 24 h. These criteria yielded 998 reference patients, who were used in both feature spaces. Median imputation and z-score standardization were fitted on the historical reference and applied unchanged to clinical patients. Each score was the mean Euclidean distance to the 10 nearest distinct reference patients. Larger distances indicated lower local representation within the corresponding feature space. Associations between these distances and subsequent 24–72-h dTIR were assessed. Patients requiring physician takeover for persistent hyperglycemia were identified separately in the plots.

### Statistics and reproducibility

Clinical comparisons used available patient-level observations. Continuous variables summarized as mean *±* SD or median (IQR) were compared using Welch’s t-tests or Mann–Whitney U tests, respectively, and categorical variables using chi-square tests. Mann–Whitney U tests were also used for CGM metrics and between-group glucose and insulin dose comparisons across three 24-h windows. Spearman’s correlations assessed associations of first-24-h mean glucose and kNN distances with subsequent 24–72-h dTIR, and of insulin dose reduction with Tmax and pump-treatment duration. Holm correction was applied to the 12 window-specific comparisons and the two kNN permutation tests. Other comparisons were exploratory, with unadjusted P values. For kNN correlations, unadjusted 95% confidence intervals were obtained from 1000 patient-level bootstrap resamples. For dose-reduction correlations, percentile 95% confidence intervals were obtained from 5000 patient-level cluster bootstrap replicates retaining all courses per sampled patient, and P values were derived from standardized-rank regression with patient-cluster-robust standard errors (CR1) and a t distribution with degrees of freedom equal to the number of patients minus one. All tests were two-sided, with significance defined as *P <*0.05 using adjusted *P* values where applicable.

### Use of generative AI

The authors used GPT-5.6 Sol through OpenAI Codex and subsequently GPT-6 Astra (OpenAI) primarily for language editing to enhance the clarity and readability of the manuscript, with additional assistance in identifying relevant literature and refining the description of results. All AI-assisted content and cited sources were reviewed and verified by the authors, who take full responsibility for the accuracy and integrity of the manuscript.

## Data availability

The datasets generated and analysed in this study are not publicly available to protect patient privacy. De-identified data supporting the findings will be available from the corresponding author upon reasonable request following publication in a peer-reviewed journal, subject to applicable ethical and institutional requirements.

## Code availability

The code used to develop and internally validate the LSTM models will be made publicly available upon publication in a peer-reviewed journal. During journal peer review, the code will be available to editors and reviewers upon request.

## Acknowledgements

This study was funded by the National Natural Science Foundation of China (82070918). The funder did not participate in study design, data collection, analysis, interpretation, manuscript preparation, or the decision to submit for publication. We thank Zhi Zhang from the Department of Endocrinology, the Affiliated Hospital of Guizhou Medical University; Xitian Zhang from the International Medical Service Center, the First Affiliated Hospital of Shantou University Medical College; Xiaomin Huang from the Affiliated Brain Hospital, Guangzhou Medical University, and Shali Liu from the First Affiliated Hospital of Sun Yat-sen University for their substantial time and meticulous work in entering historical clinical records during their postgraduate training. Their contributions provided an essential foundation for the development of this system.

## Author contributions

L.C. independently reproduced the model analyses, repeated internal validation, verified the data, prepared the figures and tables, and contributed to manuscript drafting. S.H. developed the clinical data-entry system and insulin dosing models, performed the initial internal validation, implemented the models for clinical use, and contributed to manuscript revision. Y.C. conducted the clinical trial, collected and analyzed the clinical data, and contributed to manuscript revision. Z.H. conceived and designed the study, supervised its conduct, secured funding, and drafted and revised the manuscript. All authors reviewed and approved the final manuscript.

