## Supplementary information for "A data-driven system for insulin pump dosing in type 2 diabetes: development, prospective evaluation and clinical applicability"

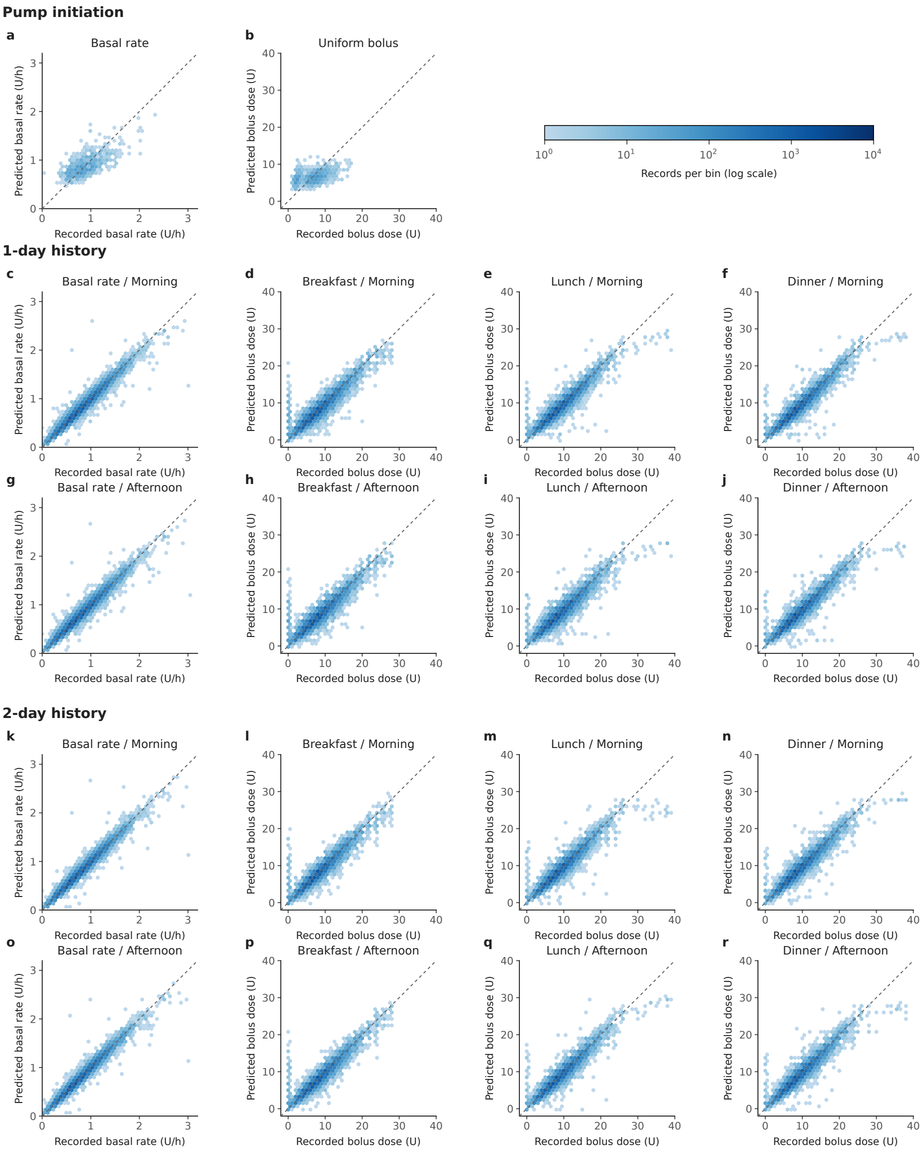


**Supplementary Figure 1  Agreement between predicted and clinician-recorded insulin doses.** Out-of-fold predictions are plotted against recorded prescriptions for pump-initiation models (a, b), one-day-history models (c–j), and two-day-history models (k–r). Dashed lines indicate identity. Color intensity represents the number of records per hexagonal bin on a logarithmic scale. Sample sizes and performance metrics are reported in Supplementary Table 1.

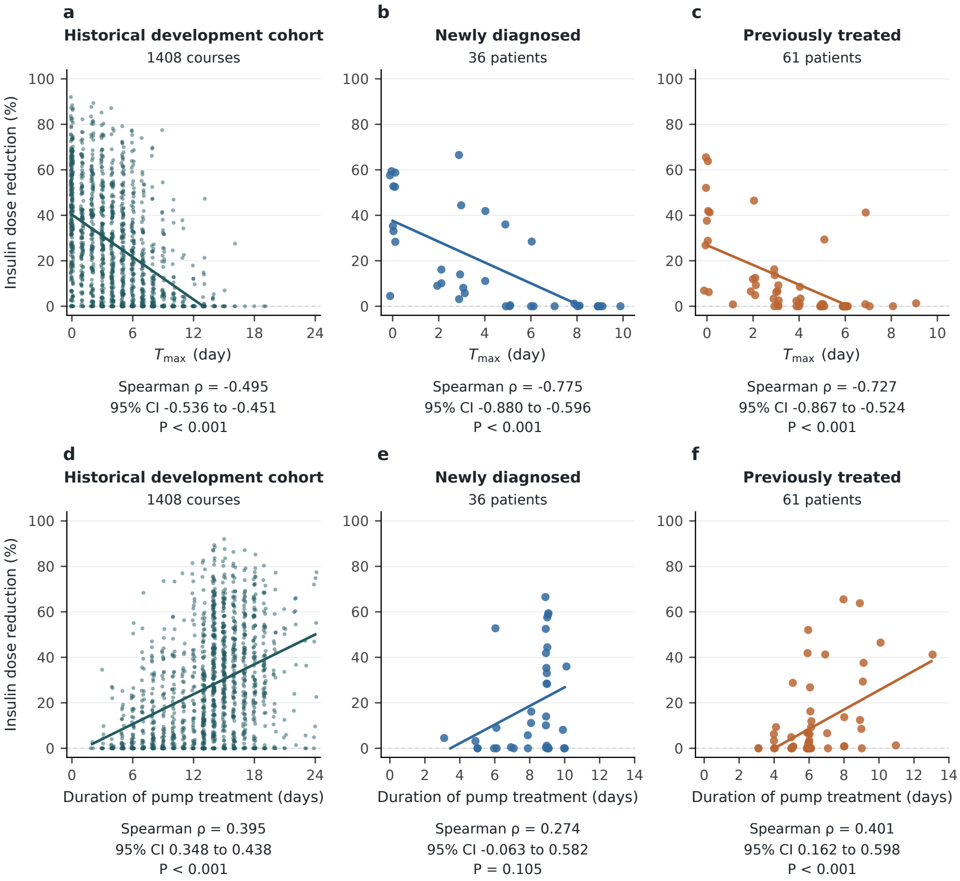


**Supplementary Figure 2 Insulin dose reduction in relation to time to maximum dose and treatment duration.** Associations of insulin dose reduction with time to maximum dose (Tmax; a–c) and treatment duration (d–f) in historical treatment courses (a, d), newly diagnosed patients (b, e), and previously treated patients (c, f). Dose reduction was calculated as the percentage decrease from the maximum prescribed daily dose to the prescribed dose on the day of pump discontinuation, with both doses calculated over a full 24 h. Points represent individual courses (historical development cohort) or patients (RCT model-directed); solid lines indicate linear fits. Spearman's correlations, 95% confidence intervals, and $P$ values are shown.

**Supplementary Table 1: Internal validation performance of the 18 stage-specific insulin dosing models.**

| Dose target (unit) | Patients n | Records n | MAE (95% CI) | RMSE (95% CI) | $R^2$ (95% CI) | Mean error |
|---|---|---|---|---|---|---|
| **Pump initiation** | | | | | | |
| Basal rate (U/h) | 1190 | 1239 | 0.150 (0.144–0.156) | 0.191 (0.181–0.199) | 0.449 (0.390–0.503) | -0.0011 |
| Uniform bolus (U) | 1182 | 1231 | 2.157 (2.066–2.247) | 2.706 (2.603–2.809) | 0.196 (0.144–0.241) | 0.0058 |
| **1-day history — Morning** | | | | | | |
| Basal rate (U/h) | 1641 | 22970 | 0.039 (0.038–0.040) | 0.061 (0.057–0.067) | 0.967 (0.961–0.971) | 0.0003 |
| Breakfast bolus (U) | 1641 | 22970 | 0.749 (0.728–0.771) | 1.211 (1.147–1.281) | 0.896 (0.883–0.908) | 0.0007 |
| Lunch bolus (U) | 1641 | 22921 | 0.678 (0.657–0.702) | 1.070 (1.000–1.146) | 0.921 (0.911–0.929) | -0.0257 |
| Dinner bolus (U) | 1641 | 22677 | 0.663 (0.644–0.683) | 1.026 (0.964–1.094) | 0.935 (0.927–0.942) | -0.0154 |
| **1-day history — Afternoon** | | | | | | |
| Basal rate (U/h) | 1641 | 22970 | 0.040 (0.039–0.041) | 0.062 (0.058–0.068) | 0.966 (0.959–0.970) | 0.0004 |
| Breakfast bolus (U) | 1641 | 22970 | 0.755 (0.733–0.778) | 1.223 (1.161–1.292) | 0.894 (0.880–0.906) | 0.0134 |
| Lunch bolus (U) | 1641 | 22921 | 0.731 (0.709–0.755) | 1.163 (1.094–1.238) | 0.906 (0.897–0.915) | -0.0471 |
| Dinner bolus (U) | 1641 | 22677 | 0.664 (0.645–0.685) | 1.035 (0.966–1.114) | 0.934 (0.925–0.941) | -0.0183 |

Continued on next page

**Supplementary Table 1 (continued)**

| **Dose target (unit)** | **Patients n** | **Records n** | **MAE (95% CI)** | **RMSE (95% CI)** | **$R^2$ (95% CI)** | **Mean error** |
|---|---|---|---|---|---|---|
| **2-day history — Morning** | | | | | | |
| Basal rate (U/h) | 1641 | 21292 | 0.036 (0.035–0.037) | 0.057 (0.051–0.063) | 0.972 (0.966–0.977) | -0.0021 |
| Breakfast bolus (U) | 1641 | 21292 | 0.694 (0.674–0.713) | 1.100 (1.044–1.160) | 0.919 (0.909–0.927) | 0.0028 |
| Lunch bolus (U) | 1641 | 21243 | 0.685 (0.664–0.710) | 1.091 (0.998–1.200) | 0.921 (0.910–0.931) | -0.0227 |
| Dinner bolus (U) | 1640 | 20995 | 0.698 (0.677–0.720) | 1.067 (1.003–1.141) | 0.933 (0.924–0.940) | -0.0333 |
| **2-day history — Afternoon** | | | | | | |
| Basal rate (U/h) | 1641 | 21292 | 0.036 (0.035–0.037) | 0.056 (0.051–0.062) | 0.973 (0.967–0.977) | -0.0019 |
| Breakfast bolus (U) | 1641 | 21292 | 0.689 (0.671–0.708) | 1.093 (1.040–1.150) | 0.920 (0.910–0.928) | 0.0181 |
| Lunch bolus (U) | 1641 | 21243 | 0.689 (0.670–0.711) | 1.066 (1.000–1.143) | 0.924 (0.916–0.932) | -0.0153 |
| Dinner bolus (U) | 1640 | 20995 | 0.700 (0.679–0.724) | 1.080 (1.009–1.162) | 0.931 (0.923–0.938) | -0.0372 |

Metrics were calculated from pooled out-of-fold predictions, with clinician-recorded prescriptions as the reference. Mean error represents predicted minus recorded dose. MAE, RMSE and mean error are expressed in the units indicated for each dose target. Patients and records may contribute to multiple tasks. MAE, mean absolute error; RMSE, root mean squared error; $\mathbf{R^2}$, coefficient of determination; CI, confidence interval.

**Supplementary Table 2 CGM-derived glycemic metrics in the model-directed group.**

| Characteristics | Newly diagnosed (n=28) | Previously treated (n=44) | *P* value |
|---|---|---|---|
| Time in range (3.9-10.0 mmol/L), TIR, % | 88.2 (78.7, 91.2) | 85.0 (68.3, 91.1) | 0.246 |
| TIR on Day 0 (%) | 66.7 (37.2, 82.4) | 42.9 (11.3, 80.0) | 0.105 |
| TIR on final analysis day (%) | 94.4 (81.6, 99.6) | 93.0 (84.8, 100.0) | 0.863 |
| Time above range, level 1 (10.0-13.9 mmol/L), TAR1, % | 7.9 (3.9, 12.3) | 12.6 (4.6, 22.9) | 0.071 |
| TAR1 on Day 0 (%) | 26.9 (10.4, 38.9) | 26.2 (0.0, 56.4) | 0.577 |
| TAR1 on final analysis day (%) | 0.0 (0.0, 3.1) | 3.1 (0.0, 10.5) | 0.057 |
| Time above range, level 2 (>13.9 mmol/L), TAR2, % | 0.1 (0.0, 1.6) | 0.7 (0.0, 6.8) | 0.077 |
| TAR2 on Day 0 (%) | 0.0 (0.0, 15.2) | 0.0 (0.0, 25.4) | 0.360 |
| TAR2 on final analysis day (%) | 0.0 (0.0, 0.0) | 0.0 (0.0, 0.0) | 0.067 |
| Time below range, level 1 (3.0-3.9 mmol/L), TBR1, % | 2.1 (0.8, 5.8) | 1.1 (0.0, 3.1) | 0.067 |
| TBR1 on Day 0 (%) | 0.0 (0.0, 0.0) | 0.0 (0.0, 0.0) | 0.842 |
| TBR1 on final analysis day (%) | 1.6 (0.0, 10.2) | 0.0 (0.0, 1.2) | 0.012 |
| Time below range, level 2 (<3.0 mmol/L), TBR2, % | 0.0 (0.0, 0.2) | 0.0 (0.0, 0.0) | 0.450 |
| TBR2 on Day 0 (%) | 0.0 (0.0, 0.0) | 0.0 (0.0, 0.0) | 0.330 |
| TBR2 on final analysis day (%) | 0.0 (0.0, 0.0) | 0.0 (0.0, 0.0) | 0.272 |
| Mean glucose (mmol/L) | 6.8 (6.2, 7.1) | 7.5 (6.4, 8.6) | 0.029 |
| Mean glucose on Day 0 (mmol/L) | 9.2 (7.5, 10.4) | 10.6 (7.6, 12.4) | 0.195 |
| Mean glucose on final analysis day (mmol/L) | 5.9 (5.3, 6.2) | 6.7 (5.9, 7.7) | 0.002 |
| Standard deviation (SD, mmol/L) | 2.1 (1.8, 2.5) | 2.2 (1.9, 3.0) | 0.129 |
| SD on Day 0 (mmol/L) | 2.1 (1.6, 2.7) | 1.9 (1.2, 2.5) | 0.305 |
| SD on final analysis day (mmol/L) | 1.4 (0.9, 1.9) | 1.7 (1.2, 2.0) | 0.171 |
| Coefficient of variance (CV, %) | 30.4 (28.0, 36.0) | 30.2 (27.5, 33.7) | 0.813 |
| CV on Day 0 (%) | 22.2 (17.9, 28.6) | 19.4 (12.0, 26.4) | 0.074 |
| CV on final analysis day (%) | 22.4 (16.5, 29.8) | 24.0 (18.1, 29.0) | 0.895 |
| Glucose management indicator (%) | 6.2 (6.0, 6.4) | 6.5 (6.1, 7.0) | 0.029 |
| Mean Amplitude of Glycemic Excursions (mmol/L) | 4.1 (3.5, 4.9) | 4.2 (3.5, 5.3) | 0.529 |

Analyses included only CGM data collected during model-directed treatment. The final analysis day is the last day of model-directed treatment, with data censored at pump discontinuation or physician takeover rather than the end of CGM monitoring. *P* values are unadjusted.
